# Sex and age-dependent alterations of drug consumption during the COVID-19 lockdown in Spain: Lessons learned for the future

**DOI:** 10.1101/2023.05.02.23289404

**Authors:** Jesús David Lorente, Anabel Forte, Javier Cuitavi, Francesc Verdú, Lucía Hipólito

**Author notes:** Corresponding author: Lucía Hipólito Dpt. Pharmacy and Pharmaceutical Technology and Parasitology Faculty of Pharmacy Avda. Vicente Andrés Estellés s/n 46100 Burjassot, Valencia (SPAIN).

## Abstract

In order to reduce the spread of COVID-19, lockdown has been one of the most implemented measures worldwide. Spain had one of the harshest lockdowns in Europe, impacting the social and psychological health of the population. The aim of this paper is to study how the lockdown has affected drug consumption patterns and the extent to which age and sex are influential factors. We have developed an online survey in which people were asked about their consumption of alcohol, marihuana, cocaine, and sedative and tranquilizers before and during the COVID-19 lockdown. Data revealed a general reduction in the consumption of all the drugs surveyed. Interestingly, when data was analysed by sex or age, we detected alterations in the consumption patterns depending on these variables that were of special relevance in the case of alcohol, marihuana and non-prescription sedatives and tranquilizers. Our data revealed a general decrease in the use of these drugs in the case of young adults, revealing that their use is strongly linked to social life, whereas the middle-aged population has experienced alterations in their consumption patterns, whereby their use has increased to daily. In addition, the use of non-prescription sedatives and tranquilizers has increased in specific populations. In conclusion, our data reveals important alterations during the lockdown in the consumption pattern of both legal and illegal drugs (sex and age dependent) in the Spanish population, and these alterations might be considered for future national strategies of preventative actions.

## Introduction

Coronavirus disease 2019 (COVID-19) has been a challenge to our health and social systems. The rapid expansion of COVID-19 entailed an avalanche of measures to prevent it from spreading across the entire World. Lockdown, together with the restriction of social activities, has been the most common measure taken, although differences in the implementation of this lockdown occurred between countries, i.e., the duration, the activities allowed outdoors, and the possibility to work at your usual workplace. Nevertheless, all these restrictions, together with social distancing, could have given rise to a large negative impact on people’s mental health due to the social distancing.

Social restrictions and psychological distress, caused by the pandemic, and their consequences in our lives have been associated with stress, depression, social isolation, suicidal tendencies and substance abuse (Holmes et al., 2020; Lamis et al., 2014; Lechner et al., 2020; Louise C. & John T., 2010). In this way, many experts have expressed their concern over the new COVID-19-derived social restrictions which, although necessary, may affect people’s mental health, promoting loneliness, anxiety, depression, and substance abuse (Galea et al., 2020; García-Álvarez et al., 2020; Holmes et al., 2020; Rehm et al., 2020). However, a myriad of alterations regarding drug intake have been observed in different populations, most probably depending on specific characteristics of the population under study or the design of the study. For example, in the case of tobacco, some researchers reported its consumption rate unaltered (Chodkiewicz et al., 2020; Stanton et al., 2020), while others reported it increased (Knell et al., 2020; Vanderbruggen et al., 2020).

Interestingly, Knell et al., 2020 showed that people over 50 years old were less likely to increase their tobacco consumption compared to people under 50, pointing to age as an important variable to take into consideration. This is further supported by studies which report similar age-related trends in drug consumption during the lockdown. For instance, several studies showed a greater alcohol consumption rate and drug use parallel to an increase in anxiety, depression and feeling of loneliness in young adults (Callinan et al., 2021; Horigian et al., 2020; Oksanen et al., 2020; Wang & Tang, 2020).

Spain underwent one of the most restrictive lockdowns in the world. It began on 15^th^ March 2020 and was extended until 21^st^ June 2020, with no possibility of engaging in any outdoor activities until May 1st when the Spanish Government allowed people to go for a walk or do exercise for a maximum of 1h per day. Undoubtedly, this long period of confinement had great repercussions on the population’s well-being, as reported in other countries. In this sense, some studies of the Spanish population show that the depressive and anxious symptoms, as well as loneliness are more common during lockdown (González-Sanguino et al., 2021; Hidalgo et al., 2020; Jacques-Avinõ et al., 2020; Losada-Baltar et al., 2021). This decline of mental well-being appeared to be more intense in the case of young adults and women (Hidalgo et al., 2020; Jacques-Avinõ et al., 2020).

Regarding legal and illegal drug use, the *Observatorio Español de las Drogas y Adicciones* (*Plan Nacional sobre Drogas*, Ministry of Health), a Spanish governmental section that provides data and statistics of drug use in Spain, has published their report on drug use in 2020 (Molina et al., 2020). Under the COVID-19 pandemic situation this report has shown important alterations of drug use that are sex and age-dependent. However, the effect of lockdown on drug use patterns compared to the pre-COVID19 situation has not been considered in this analysis.

For all the data presents above, the aim of this study is to investigate the impact of the Spanish lockdown on the consumption of most consumed drugs in Spanish population, while focusing on possible age and sex-dependent alterations. The results of this study will not only help to understand the impact of the lockdown on drug intake but will also help to analyse the drug consumption habits under an unprecedented impairment of social life that can reveal specific preventative actions to take in the current situation.

## Methods

### Participants and Recruitment

This study was reviewed and approved by the Human Research Ethics committee of the University of Valencia. The study was carried out through an online survey in the SurveyMonkey® platform, and it was published on various social media platforms (Facebook®, Twitter®, and WhatsApp®) and advertised in different newspapers, radio stations and the websites of the University of Valencia and the *Delegación de Gobierno para el Plan Nacional sobre Drogas* (DGPND, Government Delegation for the National Drugs Plan). Recruitment ran from 11th May to 30th June 2020, this period concurred with the end of the confinement in Spain. Inclusion criteria were Spanish residency and a minimum age of 18 years. From the total of 1107 surveys started, 608 were filled out completely (54.9%).

### Survey Development and Measures

The survey was adapted from the biannual EDADES survey of the DGPND of the Ministry of Health of Spain (EDADES, 2019-2020). The adaptation resulted in a 16-item survey assessing drug consumption before and after the confinement. Demographic questions included age, sex, marital status, highest level of education, and nationality. The sociodemographic characteristics of the tested population are summarized in table 1.

**Table 1.** Sociodemographic characteristics.

|  |  |  |
| --- | --- | --- |
| Gender | male | 37% |
|  | female | 63% |
| Age | 18-24 | 13.80% |
|  | 25-34 | 35.20% |
|  | 35-44 | 21.60% |
|  | 45-54 | 14.80% |
|  | 55-64 | 11.30% |
|  | > 65 | 3.30% |
| Marital status | Single | 57.20% |
|  | Married | 34.00% |
|  | Divorced | 7.60% |
|  | Widowed | 1.20% |
| Educational level | No education | 0.50% |
|  | Completed primary education | 1.80% |
|  | Completed secondary education | 8.10% |
|  | Completed further education | 19.10% |
|  | University studies | 51% |
| Nationality | Spain | 96.40% |
|  | Others | 3.60% |

Alcohol, marihuana, and cocaine consumption was assessed using self-reporting questions which describe the number of days of consumption per week during the 12 months prior to the confinement and during the lockdown.

Sedative and tranquilizers medications were assessed using self-reporting questions which analyzed whether or not people consumed sedatives and tranquilizers during the 12 months prior to the confinement and during the lockdown. Furthermore, we asked if they consumed these substances under medical prescription, and if they consumed them along with another drug (both illegal and legal).

All items used in the questions for the different drugs are presented in the tables of each drug.

### Statistical Analysis

All answers in the test were measured by qualitative variables and so are summarized using percentages and analysed through probability tests and contingency tables. Concretely, we used Chi-squared tests with a confidence level of 0.05. For the pre-post analyses, new variables were created considering if the posterior category indicates an increase, a decrease, or no change in consumption. All the analyses were performed using the programming language R (R Core Team (2021), https://www.R-project.org/).

## Results

### Alcohol

First of all, we examined alcohol consumption before the confinement. Here, we observed a significantly different consumption pattern when analysing by sex (p= 3,10E-08) or age (p= 5,00E-04) factors (Table 2). Statistical analysis revealed that men drink alcohol more days per week than women, being 3-4 days per week the most common alcohol drinking pattern in men. When analysing consumption patterns by age, the percentage of people who consume alcohol *every day* is lower in the youngest and oldest age groups. Contrastingly, age groups 45-54 and 54-65 accumulate the highest percentages of people reporting daily alcohol consumption (6 % and 13 %).

**Table 2.** Alcohol consumption pre and during lockdown.

| All population |  |  |  |  |  |  |
| --- | --- | --- | --- | --- | --- | --- |
|  | Pre |  | During |  |  |  |
| I have not consumed | 10% |  | 25% |  |  |  |
| < 1 day a week | 27% |  | 28% |  |  |  |
| Between 1-2 days | 20% |  | 21% |  |  |  |
| Between 3-4 days | 32% |  | 14% |  |  |  |
| Between 5-6 days | 7% |  | 5% |  |  |  |
| Every day | 4% |  | 7% |  |  |  |
|  | n = 608 |  | n =608 |  |  |  |
| SEX |  |  |  |  |  |  |
|  | Male |  | Female |  | P1 | P2 |
|  | Pre | During | Pre | During | p < 0.05 | p < 0.05 |
| I have not consumed | 6% | 17% | 13% | 28% | 3.10E-08 | 6.00E-04 |
| < 1 day a week | 18% | 28% | 33% | 28% |  |  |
| Between 1-2 days | 18% | 18% | 22% | 22% |  |  |
| Between 3-4 days | 41% | 19% | 26% | 11% |  |  |
| Between 5-6 days | 11% | 7% | 5% | 5% |  |  |
| Every day | 6% | 11% | 2% | 5% |  |  |
|  | n = 225 | n = 225 | n = 383 | n = 383 |  |  |

AGE
|  | 18-24 |  | 25-34 |  | 35-44 |  | 45-54 |  | 55-64 |  | > 65 |  | P1 | P2 |
| --- | --- | --- | --- | --- | --- | --- | --- | --- | --- | --- | --- | --- | --- | --- |
|  | Pre | During | Pre | During | Pre | During | Pre | During | Pre | During | Pre | During | p < 0.05 | p < 0.05 |
| I have not consumed | 11% | 43% | 6% | 21% | 10% | 20% | 12% | 20% | 14% | 22% | 25% | 30% | 5.00E-04 | 1.36E-05 |
| < 1 day a week | 36% | 26% | 27% | 34% | 3% | 27% | 19% | 20% | 26% | 20% | 25% | 30% |  |  |
| Between 1-2 days | 23% | 21% | 24% | 23% | 18% | 19% | 23% | 23% | 12% | 14% | 5% | 15% |  |  |
| Between 3-4 days | 24% | 5% | 37% | 15% | 31% | 17% | 29% | 19% | 28% | 13% | 35% | 15% |  |  |
| Between 5-6 days | 7% | 5% | 6% | 4% | 7% | 6% | 12% | 8% | 7% | 10% | 5% | 5% |  |  |
| Every day | 0% | 0% | 1% | 3% | 4% | 11% | 6% | 10% | 13% | 20% | 5% | 5% |  |  |
|  | n = 84 | n = 84 | n = 214 | n = 214 | n = 135 | n = 135 | n = 86 | n = 86 | n = 69 | n = 69 | n = 20 | n = 20 |  |  |

During lockdown, the statistical analysis revealed that alcohol intake patterns were significantly different when analysed by sex (p= 6,00E-04) or age (p= 1,36E-05), as before the confinement. Globally, both items, *I have not consumed* and *I have drunk it every day*, showed higher percentages than the pre-lockdown data when grouping by sex and age variables (Table 2). When analysing the sex variable, we observe that men developed an increase in the items *I have not consumed* and *I drink it every day*. Similarly to men, women also experienced an increase in the same items *I have not consumed alcohol* and *I drink it every day* (from 13% to 28% and from 2% to 5%, respectively) (Table 2). Furthermore, when we compared by age variable, we observed that the youngest people (18-44 years old) more frequently reported the item *I have not consumed* during confinement (43%) than before confinement (11%). In the case of the middle-aged groups (45-54 and 55-64) the items that increased more during confinement were *I have not consumed* and *I drink every day* (for detailed data see Table 2). Finally, people over 65 years old have not shown variations of their alcohol intake patterns.

Finally, to facilitate the analysis of the pre-lockdown and lockdown intake patterns we created a new variable from the data obtained. This variable represents alterations in the consumption patterns, expressed as days per week, and allows us to study if people increased, decreased, or maintained their alcohol consumption during the confinement. Table 3 summarizes the obtained results. In general, we found that 40.6% of the people that took the survey decreased their alcohol intake, 46.1% maintained it, and 13.3% increased it. Interestingly, the alcohol consumption pattern was significantly impacted by the lockdown regarding the age variable (p= 1,10E-07) while differences were not found regarding sex (p= 0.3885) (Table 3). Thus, the group between 35-44 years old declared the highest increase in alcohol consumption (19%), while the youngest groups (18-24 and 25-34 years old) reported the highest decrease (55%). Furthermore, to better describe the alterations in the alcohol consumption pattern derived from the lockdown, table 4 summarizes the distribution of people (in %) that declared drinking a number of days per week during lockdown, depending on the number of days per week that they declare consuming alcohol before lockdown. As can be observed, people declaring *I drink alcohol 5-6 days per week* before lockdown showed the highest increase in the frequency of alcohol consumption. Opposite to that, when people reported *I drink alcohol 1-2 day per week* and *I drink alcohol 3-4 day per week* before confinement experienced the highest decrease.

**Table 3.**
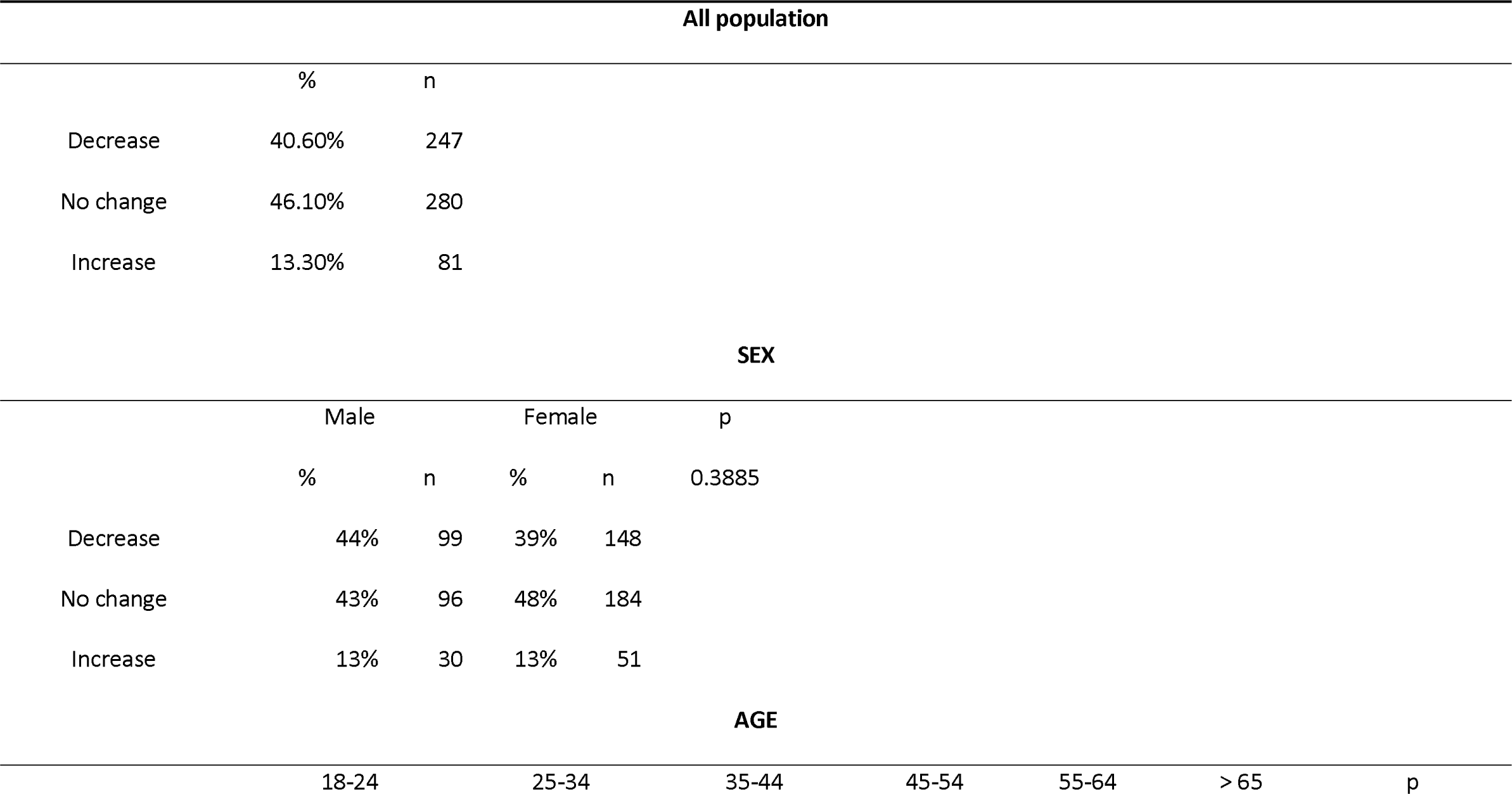
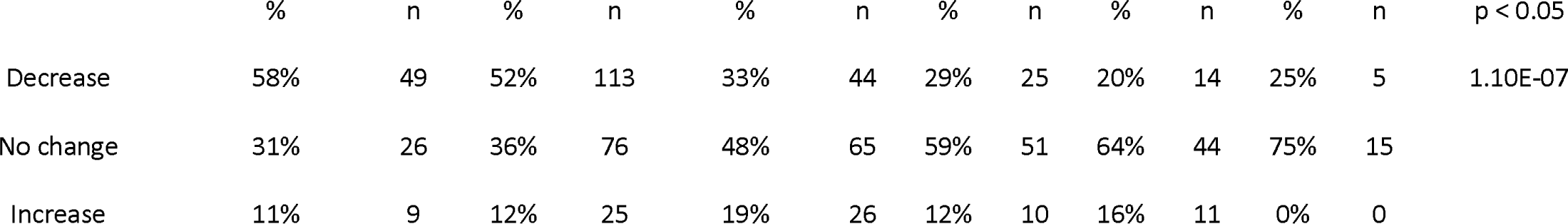
Alcohol consumption pattern changes.

**Table 4.**
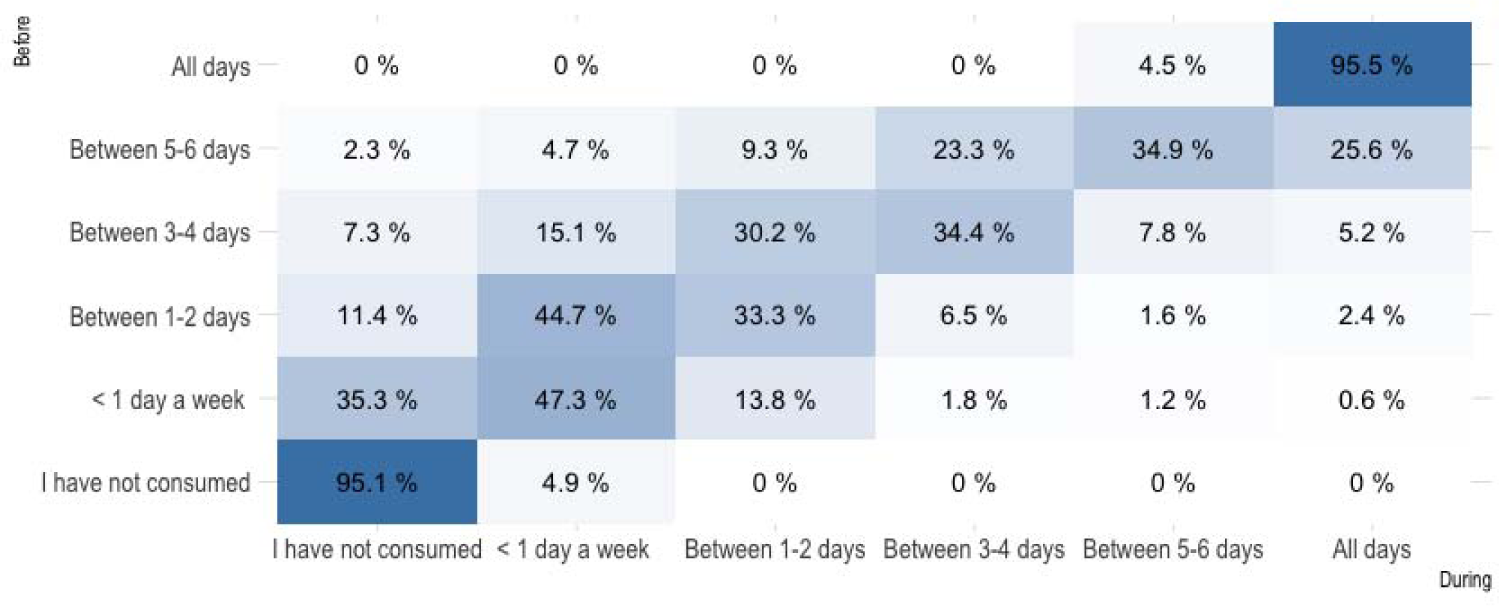
Alcohol consumption pattern changes by category.

### Sedatives and tranquilizers medication under medical prescription

21.4% of the people surveyed reported that they consumed sedatives and tranquilizers under medical prescription during the year before the confinement. This percentage was significantly higher in women (24.3%) than in men (16.4%) (p= 0.03). Furthermore, we did not find statistical differences regarding age (p= 0.29, table 5).

**Table 5.** Tranquilizers and sedatives under control pre and during lockdown.

| All population |
| --- |

|  | Pre | During |
| --- | --- | --- |
| Consume | 21.4% | 17.0% |
| No consume | 78.6% | 83.0% |
|  | n = 608 | n = 608 |

**SEX**
|  | Male |  | Female |  | P1 | P2 |
| --- | --- | --- | --- | --- | --- | --- |
|  | Pre | During | Pre | During | p < 0.05 | p < 0.05 |
| Consume | 16.4% | 11.6% | 24.3% | 20.1% | 0.03 | 0.009 |
| No consume | 83.6% | 88.4% | 75.7% | 79.9% |  |  |
|  | n = 225 | n = 225 | n = 383 | n = 383 |  |  |

**AGE**
|  | 18-24 |  | 25-34 |  | 35-44 |  | 45-54 |  | 55-64 |  | > 65 |  | P1 | P2 |
| --- | --- | --- | --- | --- | --- | --- | --- | --- | --- | --- | --- | --- | --- | --- |
|  | Pre | During | Pre | During | Pre | During | Pre | During | Pre | During | Pre | During | p = 0.29 | p = 0.67 |
| Consume | 18% | 12% | 18% | 16% | 27% | 16% | 21% | 19% | 28% | 28% | 20% | 20% |  |  |
| No consume | 82% | 88% | 82% | 84% | 73% | 82% | 79% | 81% | 72% | 72% | 80% | 80% |  |  |
|  | n = 84 | n = 84 | n = 214 | n = 214 | n = 135 | n = 135 | n = 86 | n = 86 | n = 69 | n = 69 | n = 20 | n = 20 |  |  |

During the lockdown, we observed similar results when studying by sex or by age than before lockdown. Our results showed that women continued to consume more than men (women: 20.1%, men: 11.6%; p= 0.009) without any statistical difference when analysing the results by age (p= 0.67, table 5).

Finally, we studied the evolution of the consumption of these drugs by analyzing the differences between the two periods (table 6). We observed that 90% of people did not change their consumption habits, while 7.2% and 2.8% reported that they decreased and increased their sedatives and tranquilizers intake, respectively. Nonetheless, the statistical analysis performed did not find any significant differences when comparing consumption before and during confinement by the sex (p= 0.18) or age (p= 0.64) variables.

**Table 6.** Tranquilizers and sedatives under control consumption pattern changes.

| All population |  |  |
| --- | --- | --- |
|  | % | n |
| Decrease | 7.20% | 44 |
| No change | 90.00% | 547 |
| Increase | 2.80% | 17 |
| SEX |  |  |

|  | Male |  | Female |  | p |
| --- | --- | --- | --- | --- | --- |
|  | % | n | % | n | p = 0.18 |
| Decrease | 6.20% | 14 | 7.80% | 30 |  |
| No change | 92.50% | 208 | 88.50% | 339 |  |
| Increase | 1.30% | 3 | 3.70% | 14 |  |

| AGE |  |  |  |  |  |  |  |  |  |  |  |  | p |
| --- | --- | --- | --- | --- | --- | --- | --- | --- | --- | --- | --- | --- | --- |
|  | 18-24 |  | 25-34 |  | 35-44 |  | 45-54 |  | 55-64 |  | > 65 |  | p = 0.64 |
|  | % | n | % | n | % | n | % | n | % | n | % | n |  |
| Decrease | 8.3% | 7 | 5.6% | 12 | 11.1% | 15 | 5.8% | 5 | 7.3% | 5 | 0% | 0 |  |
| No change | 89.3% | 75 | 90.7% | 194 | 86.7% | 117 | 90.7% | 78 | 91.3% | 63 | 100% | 20 |  |
| Increase | 2.4% | 2 | 3.7% | 8 | 2.2% | 3 | 3.5% | 3 | 1.4% | 1 | 0% | 0 |  |

### Sedatives and tranquilizers medication without medical prescription

6.1% of the people surveyed reported that they consumed benzodiazepines without medical prescription the year before the confinement and 6.9% during it without detectable differences regarding sex (p= 0.68), but showing differences when analysed by age (p= 0.06, table 7). Age ranges between 25 and 54 years old showed the highest percentage of non-prescription sedative consumption before lockdown (7.5%, 8.1% and 9.3% for 25-34, 35-44 and 45-54 age groups respectively). Interestingly, these age-related differences were not detected for the consumption of these drugs during lockdown (p= 0.11).

**Table 7.**
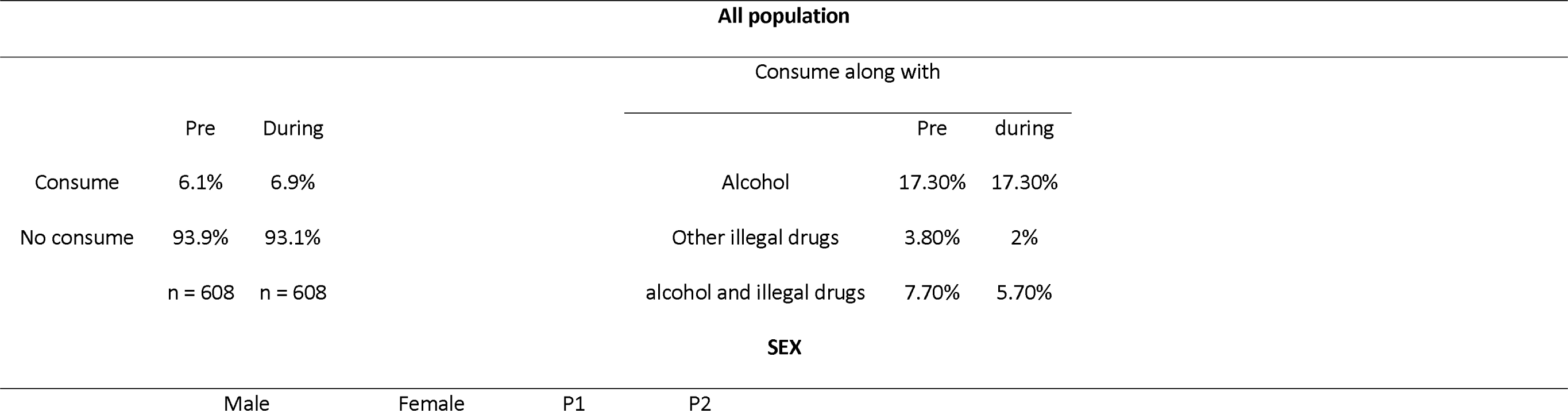
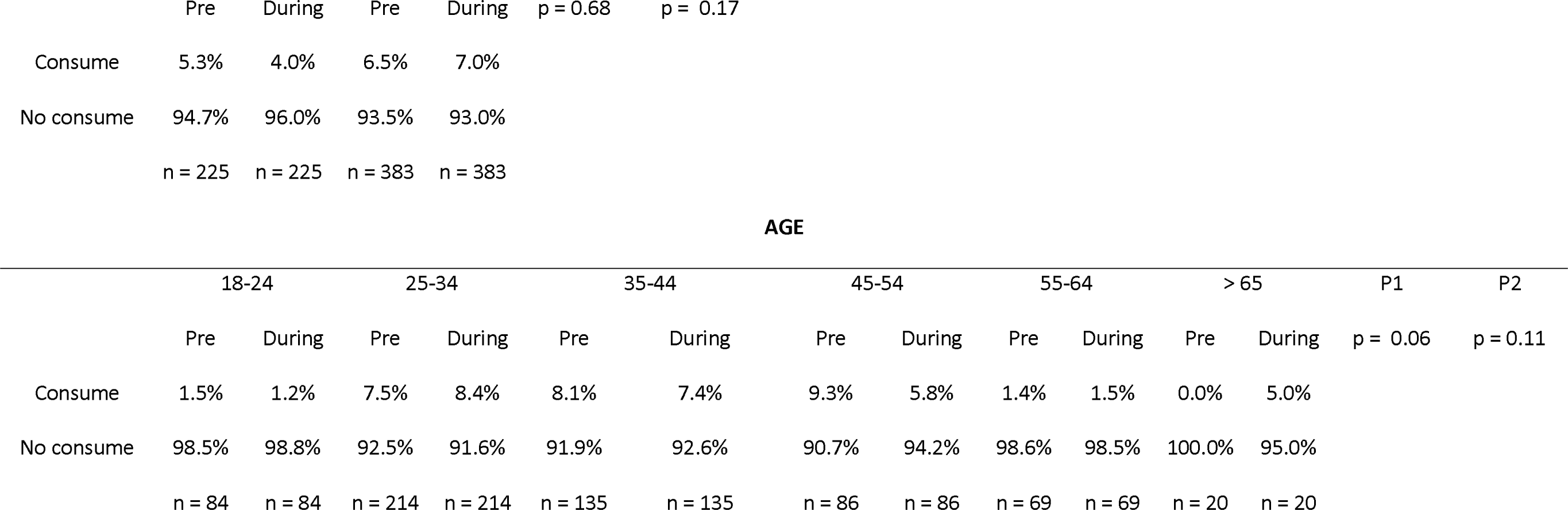
Tranquilizers and sedatives without control pre and during lockdown.

During the lockdown, we found that 94.9% of people maintained their habits, while 2.5% and 2.6% started and ceased their sedatives and tranquilizers intake. Women started to consume in a high percentage than men (3% and 1% respectively), but the statistical analysis performed did not find statistical differences when grouping individuals by sex (p= 0.38). However, we did find statistical differences in the age variable (p= 0.05). On the one hand, most of the people maintained their consumption rates, but it is interesting to note that 5% of people over 64 years old, 3.5% of the group between 45-54 years old and 4.2% of the group between 25-34 years old increased their consumption rates during lockdown. On the other hand, we observed a reduction in the intake of these substances in all group ages until 54 years old, where 7.0% of the people between 45 and 54 years old declared a reduction (Table 8).

**Table 8.**
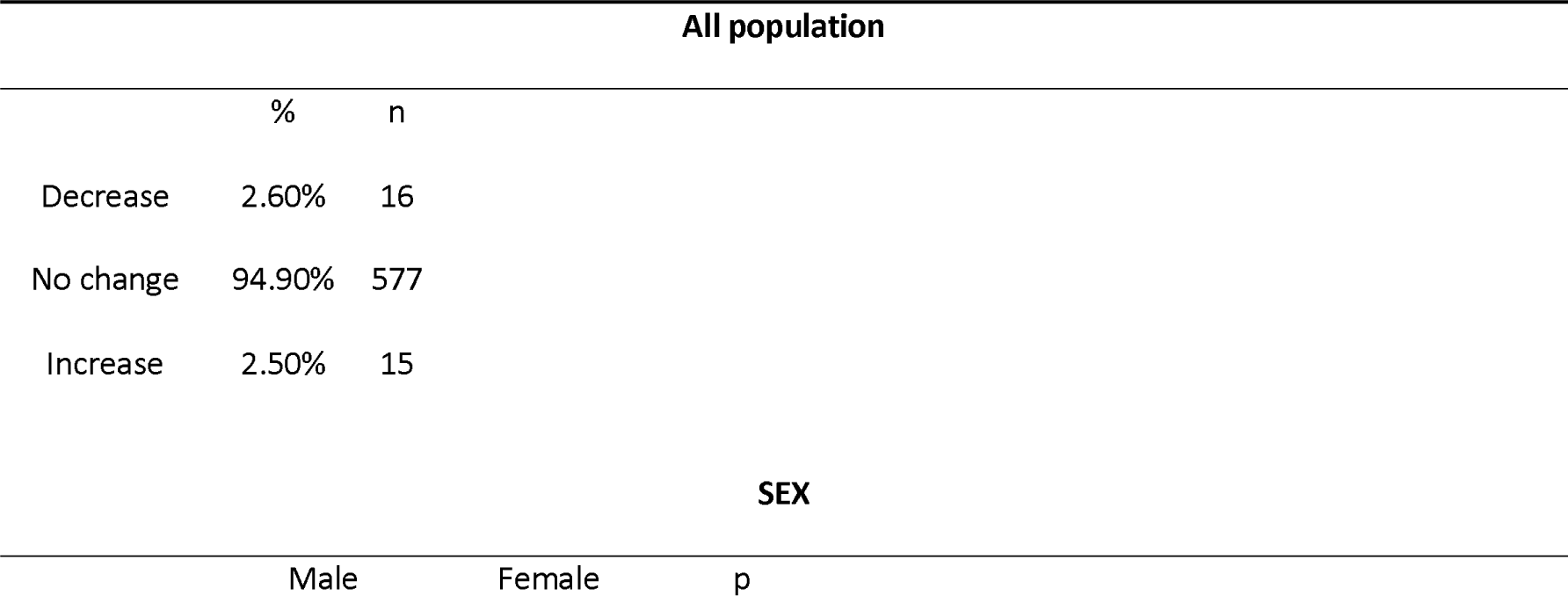
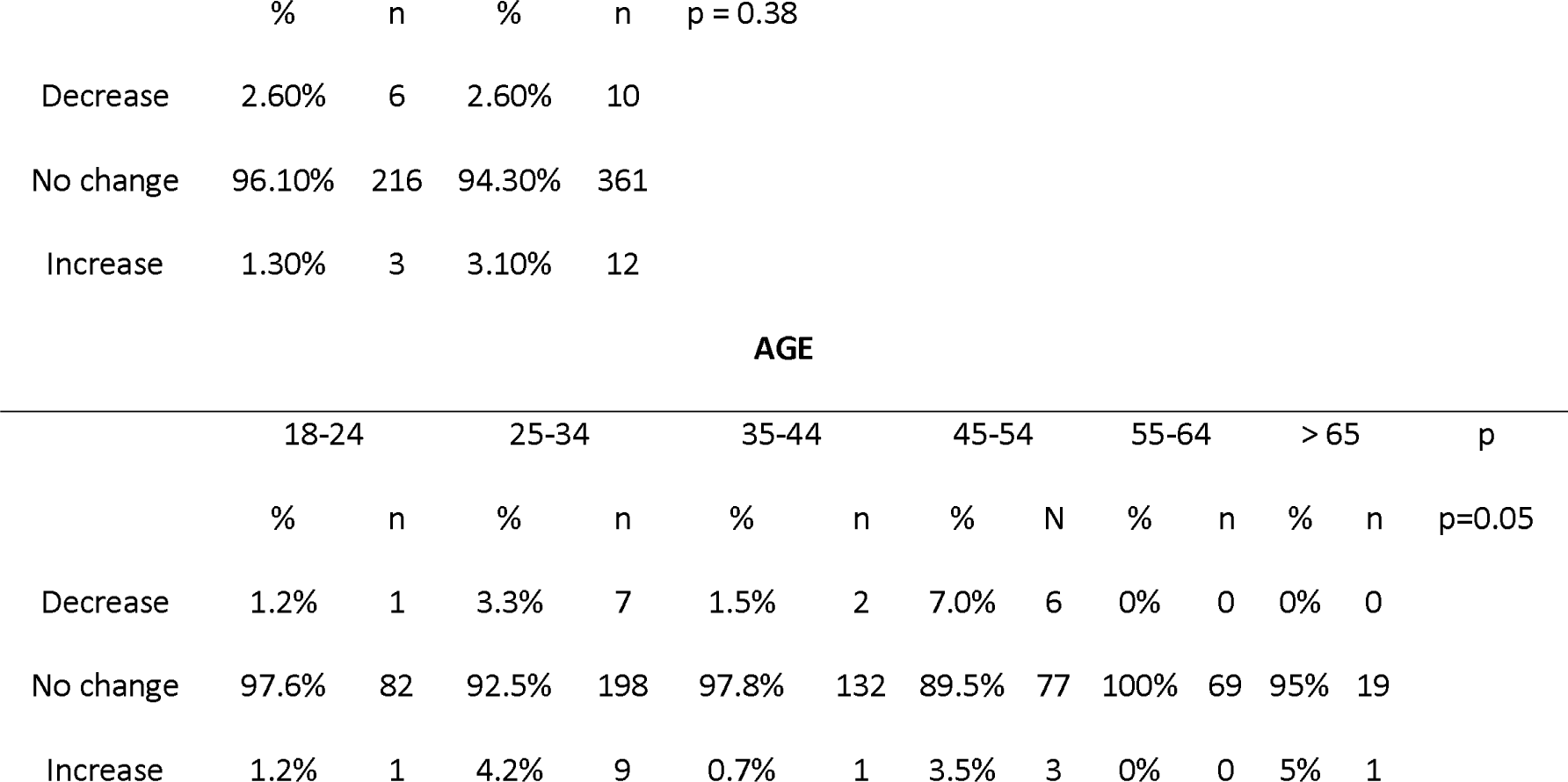
Tranquilizers and sedatives without control consumption pattern changes.

### Sedatives and tranquilizers along with other drugs

Finally, we studied the incidence of sedatives and tranquilizers intake in combination with other drugs (alcohol and/or other illegal drugs). It is important to point out that we only have 52 subjects who declared combining these drugs. Although it is a low number of people to perform a confident statistical analysis, they represent almost 9% of the surveyed sample. Before the lockdown, 17.3% of consumers reported having consumed them with alcohol, 3.8% with illegal drugs and 7.7% with both alcohol and illegal drugs at the same time. Interestingly, during the lockdown, people reported the same consumption of sedatives and tranquilizers along with alcohol, and a decrease in their combination with illegal drugs (5.7%), and with alcohol and illegal drugs at the same time (2%) (Table 7). The differences between sex and age were not analyzed because of the reduced number of subjects in this category.

### Marihuana

Firstly, we examined the consumption of marihuana before the confinement, and we discovered differences in marihuana consumption regarding the sex (p= 0.0017) and age (p= 5.00E-06) variables (Table 9). Thus, we observed that 5.3% of men reported *ever day* consumption while only 0.8% of women reported consumption on a daily basis. When investigating the consumption by age, it is interesting to note that as the age increases, the use of marihuana decreases. Indeed, 18-24 and 25-35-years old groups reported the highest percentage of *every day* (18-24: 2.4%, 25-35: 5.6%) and *several days* (18-24: 3.6%, 25-35: 3.7%) use of marihuana with the lowest percentage in the *I have not consumed* item (18-24: 69%, 25-35: 68.2%).

**Table 9.**
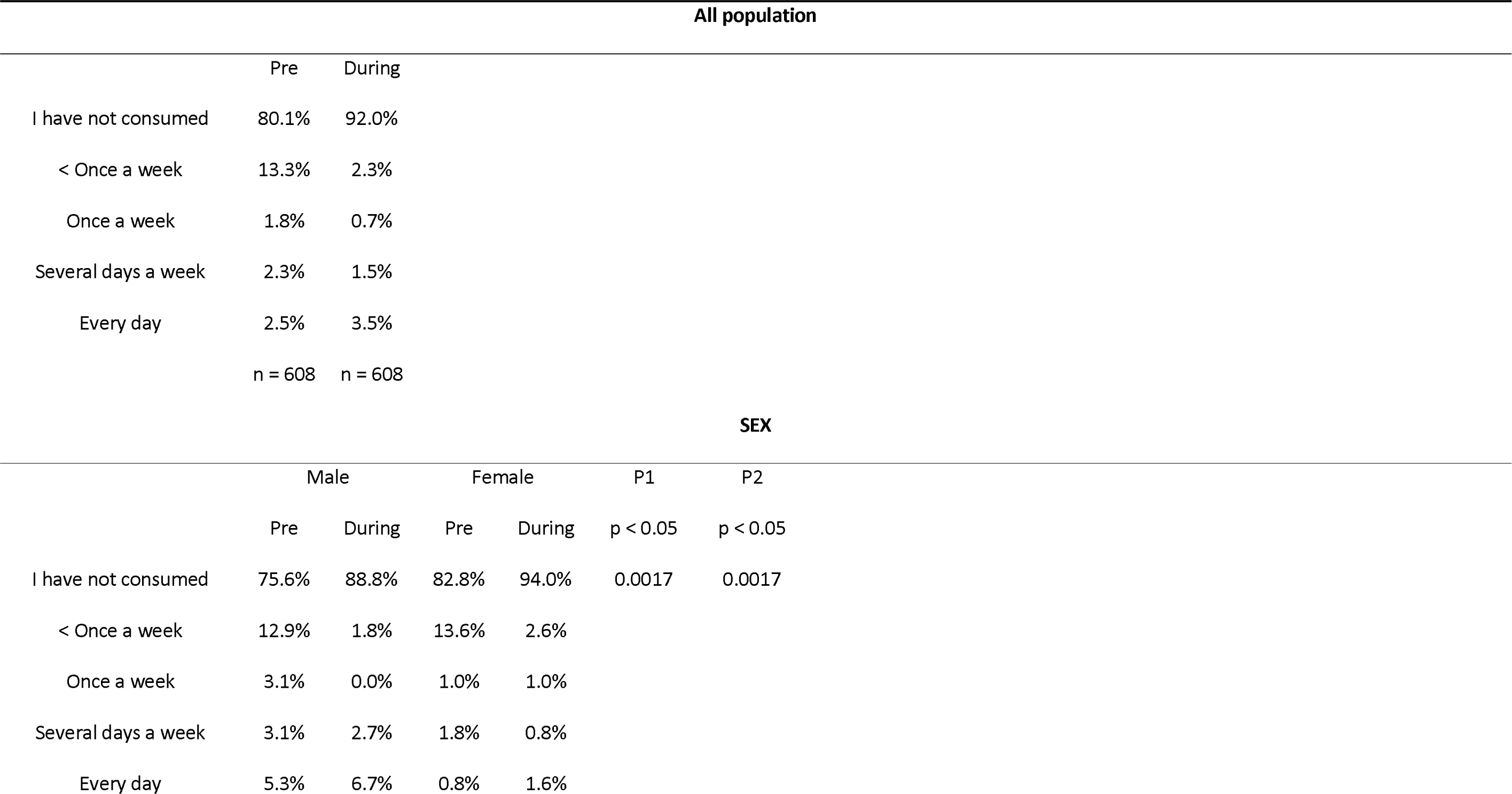
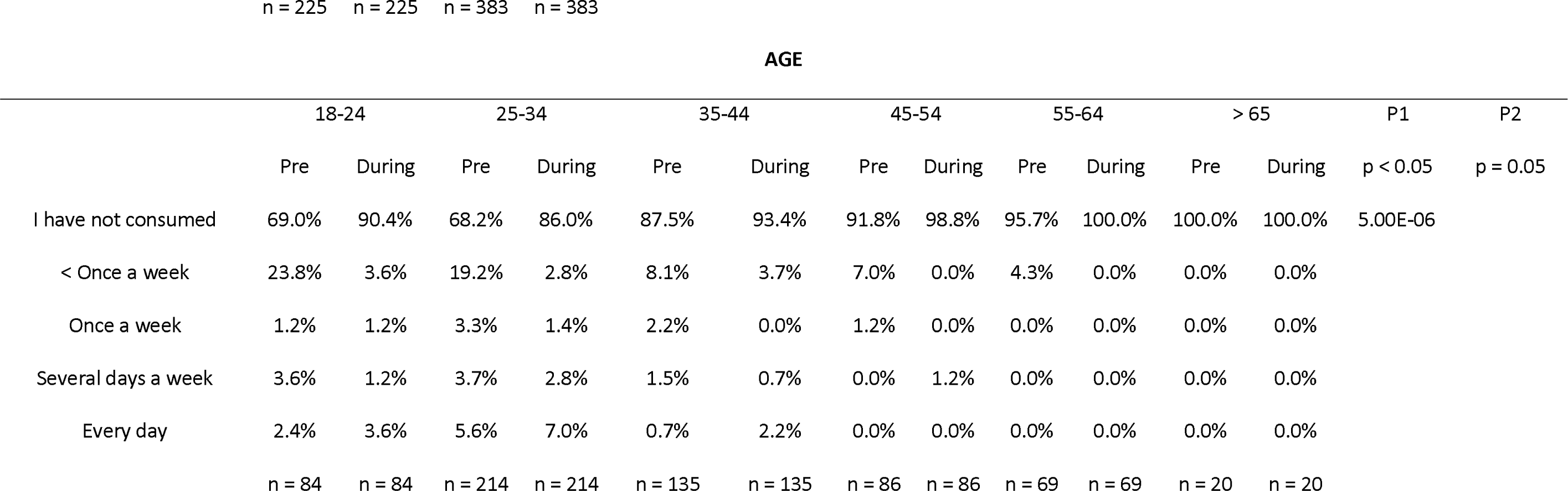
Marihuana pre and during lockdown.

During the lockdown, these statistical differences by sex (p= 0.0017) and age (p= 0.05) were maintained, although interesting alterations can be observed. Indeed, we found that the percentage of the people who do not consume was higher than before the lockdown. In fact, 92% of the people surveyed reported not having consumed marihuana during lockdown. This data indicates that 12% of the people stopped consuming marihuana during confinement when compared to the percentage that reported *I have not consumed* before confinement. Nonetheless, the surveyed people declaring *every day* consumption before the confinement also reported an increase from 2.5 to 3.5 %. Interestingly, when studying the pattern of use of marihuana grouped by ages during lockdown, we found statistical differences (p= 0.05) showing that the group between 18-24 years old and the group between 25-34 years old decreased had the most important alterations in *I have not consumed* (18-24: 69% vs 20.4%; 25-34: 68.2% vs 86%) and *every day* (18-24: 2.4% vs 3.6%; 25-34: 5.6% vs 7.0%) items (Table 9).

To further study the alterations derived from the lockdown, we also analyzed the variable created to compare marihuana use before and during lockdown. We found that most people maintained their consumption habits (84.55%), while 12.8% reduced and 2.65% increased their use of marihuana. Here, the statistical analysis did not detect sex differences (p= 0.86), but it detected interesting alterations in the pattern of use of marihuana regarding age (p= 8.00E-6). Major increases in marihuana use were observed in younger people, whereas people over 35 years old did not change their habits (Table 10). In fact, we observed that most people that reported an infrequent use of marihuana (less than once a week) reduced their consumption (82.7%), whereas people who frequently used marihuana (*once a week*), increased their consumption during lockdown (*several days a week*: 36.4% and, *every day*: 9.1%) (Table 11). This data indicates a different behaviour during lockdown depending on the previous marihuana use.

**Table 10.** Marihuana consumption pattern changes.

| All population |  |  |
| --- | --- | --- |
|  | % | n |
| Decrease | 12.80% | 78 |
| No change | 84.55% | 514 |
| Increase | 2.65% | 16 |

**SEX**
|  | Male |  | Female |  | p |
| --- | --- | --- | --- | --- | --- |
|  | % | n | % | n | p=0.86 |
| Decrease | 13.80% | 31 | 12.30% | 47 |  |
| No change | 83.50% | 188 | 85.10% | 326 |  |
| Increase | 2.70% | 6 | 2.60% | 10 |  |

**AGE**
|  | 18-24 |  | 25-34 |  | 35-44 |  | 45-54 |  | 55-64 |  | > 65 |  | p |
| --- | --- | --- | --- | --- | --- | --- | --- | --- | --- | --- | --- | --- | --- |
|  | % | n | % | n | % | n | % | n | % | n | % | n | p < 0.05 |
| Decrease | 23.8% | 20 | 18.7% | 40 | 6.7% | 9 | 7.0% | 6 | 4.3% | 3 | 0.0% | 0 | 8.00E-06 |
| No change | 70.2% | 59 | 77.6% | 166 | 91.1% | 123 | 93.0% | 80 | 95.7% | 66 | 100.0% | 20 |  |
| Increase | 6.0% | 6 | 3.7% | 8 | 2.2% | 3 | 0.0% | 0 | 0.0% | 0 | 0.0% | 0 |  |

**Table 11.**
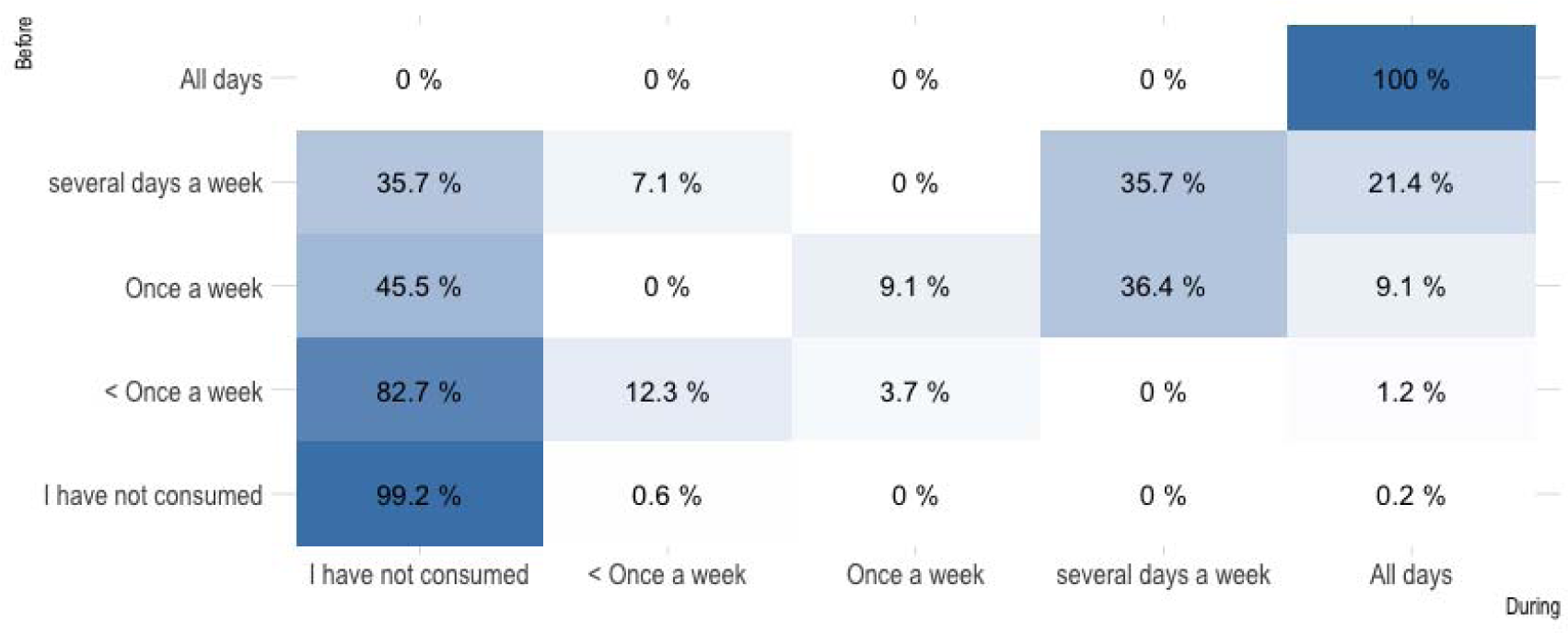
Marihuana consumption pattern changes by category.

### Cocaine

During the year before lockdown, we observed that 4.9% of the people surveyed consumed cocaine (Table 12). Furthermore, we found a higher rate of consumption in men (9.8%) and in younger people (7.9% between 25-34; 5.9% between 35-44 and less than 2% in the age groups between 45-54 and more than 65 years old), but we only detected significant differences when analyzing cocaine consumption by sex (p= 0.00023).

**Table 12.**
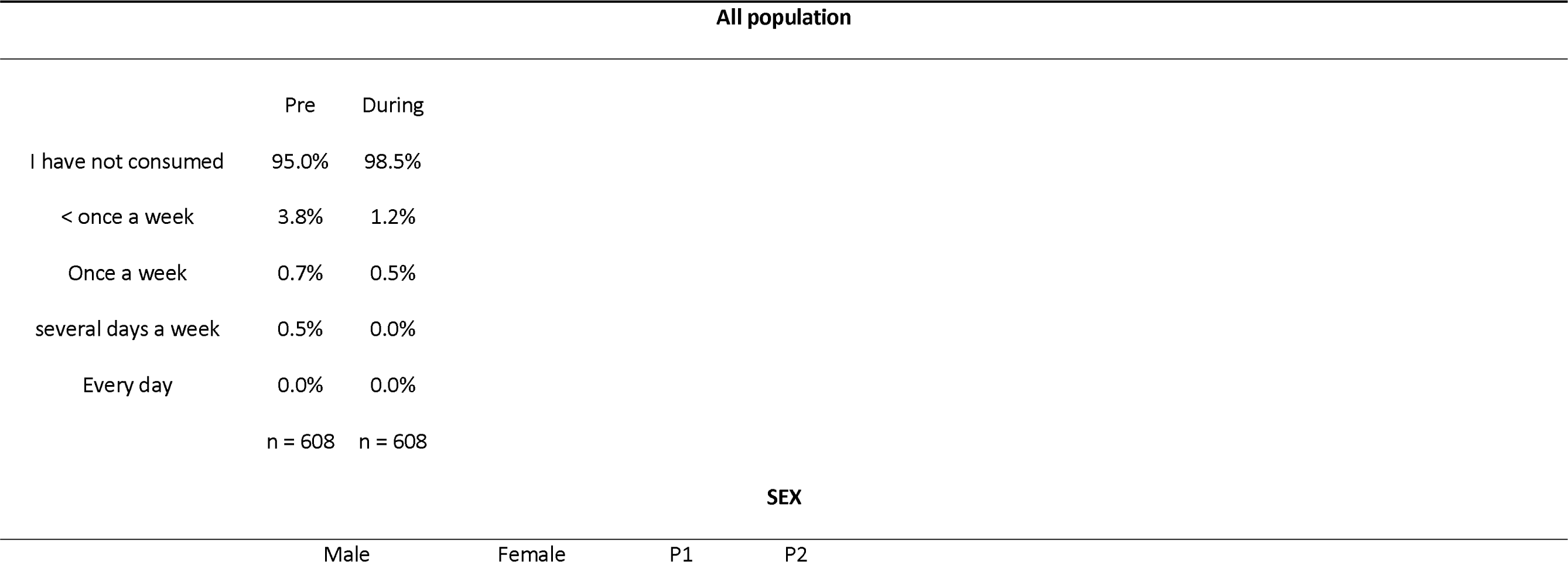
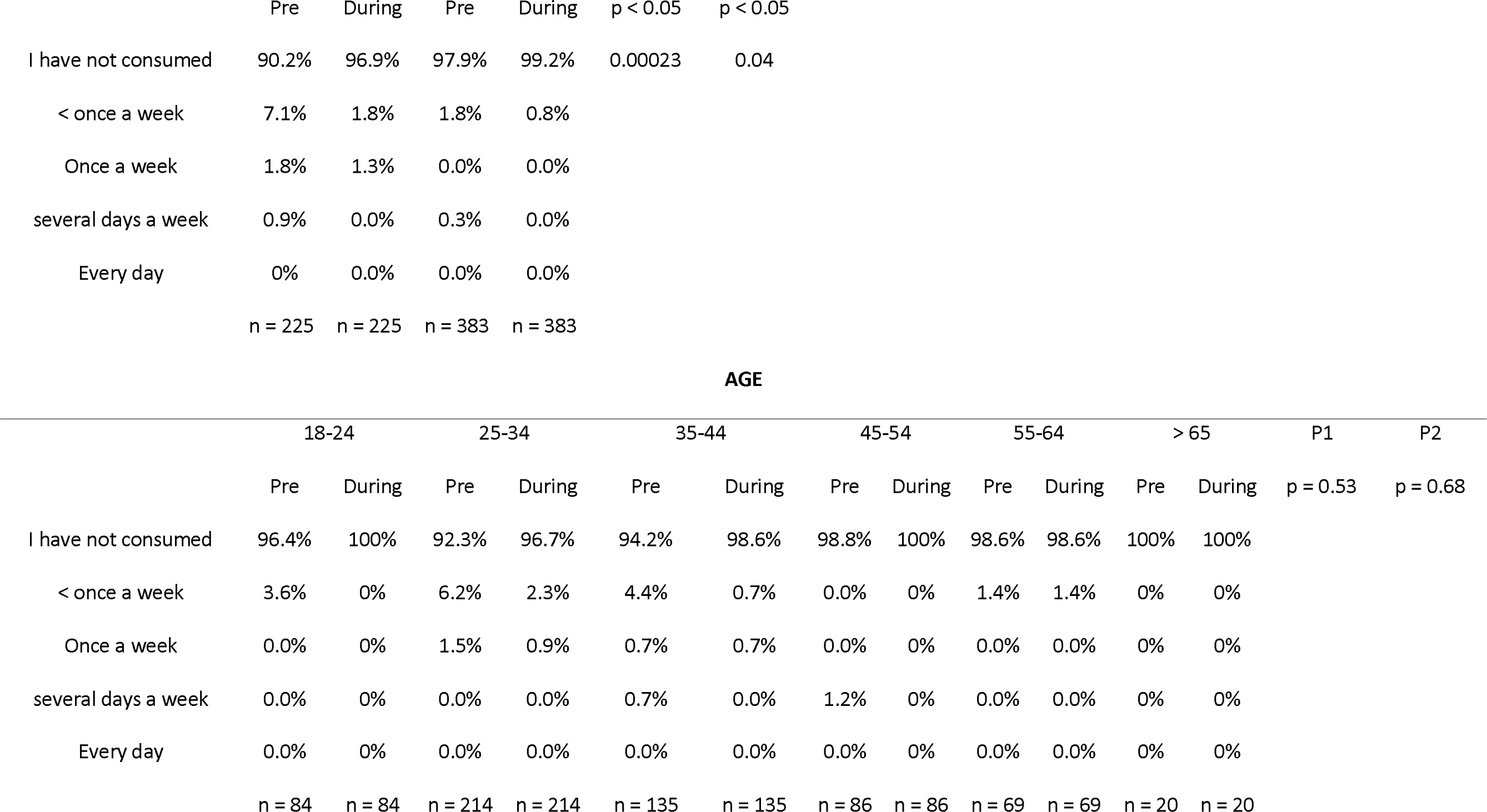
Cocaine consumption pre and during lockdown.

Remarkably, there was a general reduction in cocaine use during the lockdown (from 4.9% to1.6 %). As before lockdown the analysis of cocaine use detected differences by sex (p= 0.04) but not by age (p= 0.68). Although both sexes reduced their consumption rates, men still showed a significantly higher prevalence of cocaine consumption than women (3.1 % of men and 0.8 % of women).

Finally, table 13 shows the results obtained when comparing the consumption of cocaine before and during confinement. As can be observed, all the surveyed people maintained or reduced their intake. In fact, men reduced their consumption rates significantly compared to women (p= 0.0004), without observing alterations of the patterns of cocaine use among the different age groups (p= 0.16).

**Table 13.**
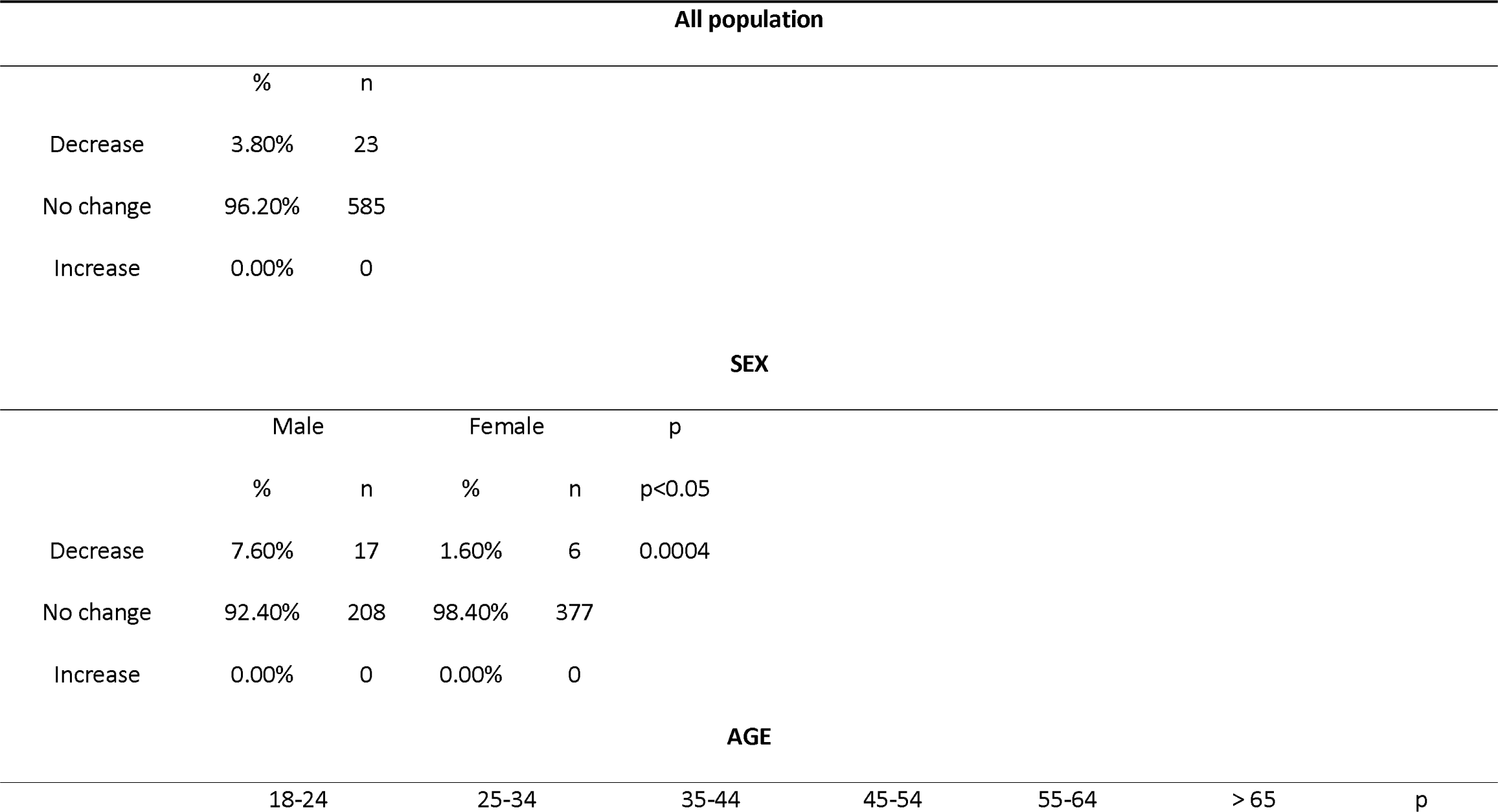
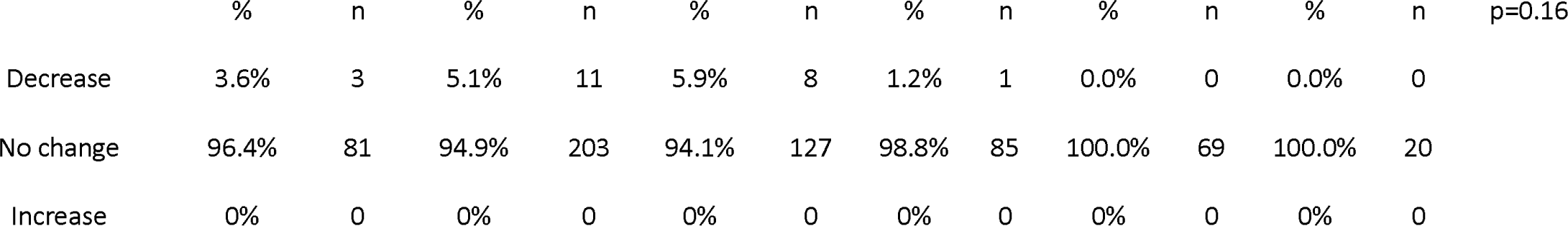
Cocaine consumption pattern changes.

### Discussion

The present study showed alterations in the drug-intake patterns before and during the confinement, which, in some cases, depend on sex or age group. These findings are not only important to understand the possible consequences of the confinement on the population, but also allowed us to detect habitual drug-intake related to social interaction that may account for political and public health actions toward the prevention of substance use disorders, especially in the youngest population.

In relation to alcohol, during the pandemic alcohol consumption increased in many different countries and populations (Ahmed et al., 2020; Callinan et al., 2021; Chodkiewicz et al., 2020; Horigian et al., 2020; Jacob et al., 2021; Lechner et al., 2020; Oksanen et al., 2020; Stanton et al., 2020). Furthermore, these high alcohol consumption rates were related to anxiety and depression symptoms, as well as negative feelings such as loneliness (Ahmed et al., 2020; Horigian et al., 2020; Lechner et al., 2020; Oksanen et al., 2020). We found that consumption was not always increased during the confinement, in fact both increases and decreases of alcohol intake were reported depending on the age variable, but not on the sex variable. Alcohol drinking patterns were statistically altered by the lockdown depending on the age. Indeed, we observed a significant decrease (53-55% of the population) in alcohol consumption in the youngest groups (18-35 years old). Very interestingly, not all the people in the group reported a reduction in alcohol consumption, a range between 11 and 19% of the population below 65 years old reported an increase in their alcohol consumption. It is interesting to note that we also observed an association between the number of days that people drank alcohol before the confinement and their consumption during it. People who used to drink 5 or 6 days per week increased their pattern of consumption to every day, and people who drank 4 days or less, decreased the number of days that they had alcoholic drinks. Two important conclusions can be drawn from this data. On one hand, we can observe that the limitation of social events impacted the young population, who stopped drinking alcohol. Lecnher et al., 2020 reported that people who used to go out to drink (i.e., not drink at home) tended to decrease their alcohol consumption during the lockdown. This data is consistent with ours, since it is well known that the young population in Spain used to drink during the weekend and that they are highly motivated to drink in a social context (Ballester Arnal & Gil Llario, 2009). This fact is of special relevance since alcohol drinking, as part of the social life of young population, not only constitutes the priming event in the use of drugs that may evolve into a drug related disorder over time, but also constitutes a key aspect for the preventative actions.

On the other hand, our data also reveals that the confinement negatively affected the middle-aged population and especially the population that already showed risky alcohol consumption behaviour, manifested by the consumption of alcohol between 5-6 days a week before the confinement. Our data is consistent with previous findings in the French population showing that regular drinkers are more prone to increasing their alcohol consumption (Constant et al., 2020). In this line, it is interesting to note that it is known that alcohol is used to affront stressful situations and/or periods, however, when the stressful situation passes, the increase in alcohol drinking may persist (Keyes et al., 2011). Finally, we cannot discard that some people increase their alcohol consumption as a substitute to other drugs intake, due to the greater difficulty in obtaining illegal drugs.

In general terms, the consumption of sedatives and tranquilizers has not been affected by the lockdown in both prescription and non-prescription use. But we found that people who took these substances without medical control changed their pattern of consumption during the lockdown, showing that certain groups increased their intake. It is notable that women consume more sedatives and tranquilizers with medical prescription than men, and these differences were maintained, with women being the ones that still showed a higher percentage in consumption of sedatives and tranquilizers during confinement. These gender differences are also observed in the impact of the confinement on mental well-being that has been previously reported (Hidalgo et al., 2020; Jacques-Avinõ et al., 2020). Interestingly, our data does not concur with other studies that reported a greater consumption of sleeping pills related with sleep disturbances that lockdown evoked in the population (Beck et al., 2021; Mandelkorn et al., 2021). Nonetheless, it is important to notice that in these studies the questionnaires included all forms of sleeping pills, including natural remedies or plant-derived compositions, whereas in our questionnaire only sedative and tranquilizer medicines were included.

On the other hand, in the case of the use of tranquilizers and sedatives without medical prescription, although the numbers show that the same percentage of people increased and decreased their consumption, statistical differences in consumption pattern alterations were observed depending on the age of the people surveyed. Indeed, the higher increases were observed in young adults and people older than 65 years old. Although we cannot analyse the causes of these specific pattern alterations, it may indicate the special vulnerability of these two age groups to suffer from the negative emotional consequences of the social isolation or work-related problems caused by the lockdown. In addition to this, it is important to highlight that the consumption of these drugs without medical prescription is illegal in Spain and the sources of these medicines, in the restrictive Spanish lockdown, makes access to these drugs difficult. One possible source is the availability of these medicines in Spanish homes because of old prescriptions that have not been retired from the prescription system or over prescription to other members of the family. Data from the Spanish Agency of Medicines (AEMPS, *Agencia Española de los Medicamentos y Productos Sanitarios*) and from the 2021 Report from the International Narcotics Control Board (INCB) show that the use of these medicines has been increasing in Spain over years, with Spain being the country with the highest number of defined daily doses per 1000 habitants in 2021 (AEMPS, 2021; INCB, 2021). This data is highlighting that the prescription of sedatives and tranquilizers and/or the deficit in their prescription control might be carefully studied.

Regarding marihuana consumption, we reported a significant reduction in its use. It is interesting that men reported a higher consumption than women before and during the lockdown since no differences between sex in the comparative analysis of pre-lockdown versus lockdown timing were observed. In previously published studies in France and Belgium, very similar data was reported (Rolland et al., 2020; Vanderbruggen et al., 2020). Curiously, surveys done in Nederland, where marihuana is legal, have revealed an increase in the amount and the patterns of consumption during the lockdown (Cousijn et al., 2021; van Laar et al., 2020). The discrepancy in the patterns between countries can be interpreted as changes of motivation for consuming the drug during the confinement depending on the accessibility of said drug. It is important to note that we found a general decrease in marihuana consumption during the confinement that was more pronounced in young people. This could also be, as alcohol consumption, related to the habits of young people in consuming marihuana during their social life. Nonetheless, it is also interesting to note that young people were also the ones that showed the highest rates of increase in marihuana use. Finally, data regarding alterations in the pattern of consumption depending on previous use; revealed that people not changing or increasing their patterns of marihuana consumption were those who took it regularly. This data confirms something that is well known in the field of drug use disorders: in order to develop a drug use disorder, people who consume more regularly are more prone to repeat the use and lose control of their intake, especially in stressful situations (Koob & Volkow, 2010).

Finally, we show that, overall, cocaine consumption was dramatically reduced probably because the impossibility of going out to obtain this drug. Interestingly, this did not happen with marihuana consumption where most people maintained their habits, most probably because of its availability which, in some cases, can even come from self-production.

It is interesting to note that this study has limitations. First of all, the relatively small sample size (n= 608) that has limited the ability to perform statistics in some of the analysis (i.e. the use of sedatives and tranquilizers along with other drugs). Moreover, the data was only obtained during the confinement period, and it is limited to the beginning of the COVID-19 crisis. As a consequence of the timeline of data recollection, a potential recall bias should be taken into consideration because we also asked about their consumption during the 12 months prior to the confinement.

Our findings indicate that the use of both legal and illegal drugs might be monitored during stressful situations in specific population depending on age and sex. But even more interestingly, our data reveals that the drug consumption habits of young people are conditioned to social interaction. Indeed, this group stopped using drugs, especially alcohol, cocaine and marihuana, to a great extent in the absence of the possible social life with their peers. Nonetheless, it is also interesting to remark that young adults were also the ones reporting a higher increase in *every day* consumption of marihuana. In our opinion, the confinement period has provided us the opportunity to conduct a very interesting experiment in restricted social interaction conditions that, until now, we have not been able to carry out with the Spanish population. This opportunity has uncovered specific characteristics of the modern habits of drug consumption in the young Spanish population that might be taken into account for preventative and public health actions in the future. Nonetheless, the situation of middle-aged people is observed to be different, especially in the case of alcohol, which is the more accessible drug. In this population, where alcohol consumption has been already established in their lifetime, this stressful event has been critical to induce a higher intake of the drug affecting, above all, consumption patterns. Finally, it is also interesting to highlight the problems surrounding the consumption of non-prescription sedatives and tranquilizers. Although the data does not reveal big alterations in the percentage of its use, the diversion of the medical prescriptions for the use of these drugs in specific populations (depending also on sex and age variables) should be a critical point to include in preventative actions. Indeed, the development of healthy entertainment and social life, together with specific actions towards the rational use of tranquilizers, sedatives and opioid medicines, are two of the milestones to achieve in the current Drug Action Plan of the (DGPNSD).

## Data Availability

All data produced in the present study are available upon reasonable request to the authors

## Acknowledgements

This study has been supported Spanish Ministerio de Sanidad, Delegación del Gobierno para el Plan Nacional sobre Drogas PNSD2019I038 (L.H.). We thank Mr. Darren We also thank Ms. Pilar Laso for her task as grant manager. We would also like to thank Mr. Darren Robinson for English Proofreading.

